# Online prescribing for sexually transmitted infections – what’s on offer? An evaluation of internet pharmacies based in the United Kingdom

**DOI:** 10.1101/2021.11.27.21266931

**Authors:** Elizabeth Okecha, Emily Boardman, Saleha Patel, Emile Morgan

## Abstract

**Background:** Online pharmacies offer an alternative approach for patients to manage their sexual health. Our aim was to determine the type of antimicrobials sold as treatment for sexually transmitted infections (STIs) by UK internet pharmacies and if providers were adhering to national guidelines.

**Methods:** A search engine results page (SERP) generated a list of registered UK online pharmacies offering treatment for the following infections: *Chlamydia trachomatis, Neisseria gonorrhoeae, Herpes simplex and Trichomonas vaginalis*. An initial audit in 2017 benchmarked each provider against the British Association of Sexual Health & HIV (BASHH guidelines. Results were fed back to each provider before re-audit in 2020. Websites selling antibiotics for non-gonococcal urethritis (NGU) and *Mycoplasma genitalium* were included at re-audit.

**Results:** There were 30 pharmacies identified in 2017 of which, five were excluded. Treatment could be obtained for *Neisseria gonorrhoeae* from five pharmacies without providing a culture result; three (60%) pharmacies sold BASHH approved antibiotics for *Neisseria gonorrhoeae*. All 25 pharmacies sold *Chlamydia trachomatis* treatment; 22 (88%) offered first line treatment options but no website assessed for proctitis. *Herpes simplex* treatment was sold on 22 websites of which, 13 (59%) offered treatment recommended by BASHH. *Trichomonas vaginalis* treatment was sold by four websites in line with BASHH. Results at re-audit showed an improvement in standards, although advice before, during and after treatment remained variable.

**Discussion:** Our work has allowed us to engage with providers to improve prescribing within the UK online pharmacy industry. However, tougher regulation is needed in order to embed sustainable change for patients who choose to access treatment online.

## Introduction

Online healthcare has seen significant levels of growth within the last decade. An analysis of the market predicts that by 2025 the global market in online pharmacies is predicted to be worth $131 billion.[1] In the United Kingdom (UK), a online pharmacy can be owned by Directors with no clinical expertise. However, a registered Pharmacist must be employed by the company for a pharmacy to obtain registration with the General Pharmaceutical Council.[2] Some companies employ Doctors to review patient information/prescribe; many have no qualifications in Sexual Health.

In 2016, the British Association for Sexual Health and HIV (BASHH) recognised a growing trend in online pharmacies selling treatment for sexually transmitted infections (STIs).[3] Around the same time, we started to see patients presenting to our service that had purchased treatment online. This prompted us to investigate the types of treatment available via the internet. To our knowledge, this is the first study of its kind to investigate online STI prescribing in the UK.

## Methods

Using the Google™ UK search term “online treatment for STIs” we were able to generate a list of pharmacies selling treatment for *Chlamydia trachomatis, Neisseria gonorrhoeae, Herpes simplex and Trichomonas vaginalis. Treponema pallidum* treatment was not available. Non-UK sites and providers requesting credit card/mobile phone verification prior to assessment were excluded. A mystery shopper audit was performed between January/February 2017 using the following standards:

- Evidence of self-assessment prior to purchase
- Treatment sold to over 18s
- Written information in line with BASHH patient information (PILS)
- Indications for treatment/antimicrobials in line with BASHH

Each provider was given feedback and time to implement recommendations. Re-audit was performed between March/April 2020; procuring a research phone expanded our inclusion criteria. Pharmacies offering treatment for non-gonococcal urethritis (NGU) and *Mycoplasma genitalium* were included.

## Results

### 2017 audit

In total 30 websites were identified, 25 met inclusion criteria. We excluded 5 websites (website upgrade/verification required with a mobile phone and/or credit card).

#### Neisseria gonorrhoeae

Five (20%) pharmacies sold treatment for *Neisseria gonorrhoeae*. One (20%) offered first line treatment [4] administered at a Nurse led clinic. Two (40%) sold alternative treatment in line with BASHH guidelines.[4] The remaining two (40%) websites did not comply with BASHH (Cefixime 400mg stat and Cefixime 400mg stat/Azithromycin 500mg respectively). No websites requested evidence of *Neisseria gonorrhoeae* culture. Two (40%) websites advised patients to abstain from sex during treatment, three (60%) advised a test of cure and partner notification. All five (100%) offered written information.

#### Chlamydia trachomatis

Treatment was available on 25 (100%) websites. Of these, 22 (88%) websites offered treatment in line with BASHH guidance.[5] Two (8%) websites sold Azithromycin outside of guidance (two gram stat or 500mg stat followed by four days of 250mg daily). One (4%) website incorrectly dosed Azithromycin and offered Ciprofloxacin 1.5 gram stat which was not BASHH approved.

Written information was available on 21 (84%) websites; three (12%) providers inappropriately advised test of cure before 14 days of treatment. Partner notification was advised on 18 (72%) websites and 16 (64%) websites advised patients to abstain from sexual contact during treatment. One (4%) provider incorrectly advised patients they could have sex during treatment if condoms used. No website assessed patients for rectal symptoms or had information on *Lymphogranuloma venereum* (LGV).

#### Herpes simplex

22 websites sold treatment. Three (14%) pharmacies mandated a photograph of skin lesions or test results before prescribing treatment. Six (27%) sold treatment without assessing if the patient had been diagnosed by a clinician. 12 (54%) sold treatment based on self-declaration of a previous diagnosis and one (5%) gave the option to upload a photo, however this could be bypassed.

13 (59%) pharmacies offered BASHH recommended treatment for first episode/recurrence. [6] Six (27%) pharmacies sold alternative treatment recommended by BASHH. One (5%) pharmacy sold Famciclovir at the correct dose however Valaciclovir was found to be sold incorrectly as 500mg once a day for five days. One (5%) pharmacy prescribed Valaciclovir correctly but sold topical Aciclovir against guidelines. One (5%) pharmacy sold Valaciclovir as a 10 day course, double the recommended treatment duration.[6]

Written information was available on 19 (86%) websites and only seven (32%) advised patients to inform new partners. One (5%) website advised patients that partners should be tested for *Herpes simplex* without clarifying testing was only indicated if partners had symptoms. In total 11 (50%) pharmacies had clear instructions to avoid sex until symptom free. Two (9%) websites had instructions to avoid sex but information was not easily accessible. One (5%) pharmacy advised against sexual contact until seven days after treatment and eight (36%) had no written information on the avoidance of sexual contact until symptom resolution.

#### Trichomonas vaginalis

Four websites were identified that sold *Trichomonas vaginalis* treatment. All four (100%) offered Metronidazole 400mg twice daily for seven days aligning with BASHH recommended first line options.[7] Additionally all websites provided customers with written information prior to purchase. All websites advised patients to avoid alcohol during treatment; no website recommended abstinence from sexual contact during treatment. Partner notification was discussed on two websites (50%).

### 2020 Re-audit

In total, 31 websites were identified, of which four were new. Three pharmacies had ceased trading and one pharmacy was again excluded for credit card verification prior to assessment. Of the 27 pharmacies included, all 27 (100%) achieved the baseline assessment standard (see table 1)

**Table 1.** Information gathered by websites prior to purchase

| Table 1 – Information gathered by websites prior to purchase |  |  |
| --- | --- | --- |
| Baseline assessment | 2017<br>n=25 | 2020<br>n=27 |
| Past medical history | 24 (96%) | 27(100%) |
| Drug history | 22 (88%) | 27(100%) |
| Allergy status | 24 (96%) | 27(100%) |
| Treatment for over 18s | 25 (100%) | 27(100%) |
| Pregnancy risk/breastfeeding | 19 (76%) | 27(100%) |

#### Neisseria gonorrhoeae

Of the five websites identified in 2017, two (40%) had withdrawn treatment from sale and one (20%) pharmacy only sold treatment to patients able to provide evidence of a valid prescription. Of the two remaining websites, both signposted customers to obtain treatment via a Sexual Health clinic although patients were still able to purchase oral treatment through both pharmacies. Again, treatment was accessible without providing a culture result. Both pharmacies prescribed treatment in line with BASHH oral alternatives, provided written information, advised abstinence during treatment and recommended partner notification. Test of cure was only recommended by one (50%) pharmacy.[8]

Two new providers sold *Neisseria gonorrhoeae* treatment. Both assessed patients for treatment indications with one pharmacy only supplying antibiotics to customers who were with evidence of a testing. Only one (50%) provider signposted patients to Sexual Health before allowing access to the self-assessment page. Oral alternatives sold on both websites were in line with BASHH alternatives although evidence of a *Neisseria gonorrhoeae* culture result was not required to purchase antibiotics. Both websites provided written information and recommended partner notification. Only one (50%) website advised users to abstain from sexual contact during treatment and recommended a test of cure two weeks post-treatment.

#### Chlamydia trachomatis

Treatment was sold by 24 providers (20 from the initial audit, four were new). Of the 20 providers from our baseline audit, all 20 (100%) assessed patients for treatment indications. 14 pharmacies (70%) assessed patients for rectal symptoms; 19 (95%) sold first line BASHH recommended treatment for *Chlamydia trachomatis* [9], although four (21%) also sold the alternative BASHH treatment option of Azithromycin, but at incorrect doses (one gram stat or 500mg stat followed by 250mg for four days). Written information on *Chlamydia trachomatis* was available on 18 (90%) websites and 17 (85%) websites advised patients to abstain from sex and notify sexual contacts. Only 6 (30%) advised users under 25 to access repeat testing 3 months after purchasing treatment.

Of the four new providers, all assessed patients for treatment indications. Three (75%) routinely enquired about rectal symptoms and all four (100%) sold Doxycycline in line with current BASHH standards. Alternative antibiotic options were available on two (50%) pharmacies; these were not in line with BASHH alternative options. One provider sold Azithromycin 1 gram stat while the other offered Oxytetracycline and Lymecycline.

Written information and advice to abstain from sex during treatment was available, although advice on partner notification only appeared on three (75%) websites. None of the new websites appeared to promote *Chlamydia trachomatis* testing three months post-treatment for under 25s.

#### Herpes simplex

Treatment was available from 23 providers (19 from previous audit, four new). Of the 19 original providers, 14 (74%) advised patients to avoid sex when lesions were present. Three (16%) required users to provide evidence of a photograph of skin lesions or positive swab result. Written information was on all 19 (100%) of the websites from our original study although advice to disclose diagnosis to new partners was only on eight (42%) websites. 17 (89%) websites supplied treatment to patients experiencing their first episode of *Herpes Simplex*. Although most of these websites were found to offer treatment options in line with BASHH guidelines[7], one (5%) provider was found to be selling treatment at a lower dose than recommended (Valaciclovir 500mg once daily) for first episode/recurrence.

Of the four new providers selling *Herpes simplex* treatment, partner disclosure was only recommended on one (25%) website. All four (100%) pharmacies advised patient to abstain from sex when lesions present but no pharmacy asked for evidence of a swab or photograph prior to purchase. Three (75%) websites offered antivirals in line with BASHH first episode/recurrence. However, one (25%) sold Famciclovir 250mg three times a day for five days for recurrent symptoms which was not a BASHH approved.

#### Trichomonas vaginalis

No new providers were found to be selling treatment. Five websites from our original audit were still selling Metronidazole at correct doses. Only two (40%) assessed for symptoms prior to sale and one (20%) required patients to self-certify that they or a partner had tested positive for infection. Each provider clearly informed patients to use low doses during pregnancy. Advice to abstain from alcohol as well as partner notification was present on four (80%) websites and abstaining from sex was mentioned on three (60%) websites.

#### Mycoplasma genitalium

Four websites sold treatment. Only two (50%) pharmacies sold antibiotics in line with current first line BASHH treatment options [10]. The remainder sold a week of Doxycycline monotherapy or the correct combination of Doxycycline/Azithromycin but supplied Azithromycin at incorrect dosage (one gram stat). One website (25%) required evidence of a positive test prior to payment and recommended test of cure, although this was earlier than the recommended time interval. Nevertheless, written information was available on all websites. Partner notification was only clearly stated on three (75%) websites.

### NGU

Three websites sold treatment. One (33%) required a valid prescription before online assessment so we were unable to evaluate this provider. Of the remaining two, one (50%) did not require self-declaration of an NGU diagnosis to purchase antibiotics /evidence microscopy. The other website did ask if the patient had attended Sexual Health prior to selling treatment but did not mandate microscopy results. Treatment sold on one (50%) website was in line with BASHH first line recommended guidance.[11] One (50%) website was selling Azithromycin one gram stat for NGU which was not in line with current practice. Both providers recommended abstaining from sex for 14 days from the start of treatment and written information on NGU and partner notification was deemed to be clear. Nevertheless, only one (50%) pharmacy recommended testing for *Chlamydia trachomatis and Neisseria gonorrhoeae* and neither one discussed *Mycoplasma genitalium* testing.

## Discussion

Our methodology was limited to reviewing website information prior to purchase due to the cost implications of purchasing treatment from each website. Additional information may have been available to patients after purchase; based on responses received from pharmacies, we don’t believe that this is likely to have been the case.

Our exclusion criteria for the initial audit included websites that required mobile phone verification leading to a reduction in the number of websites we were able to include. The addition of a research phone in 2020 helped us to identify a wider breadth of websites at the time of re-audit. Nevertheless, websites excluded due to credit card verification may have been prescribing outside of guidelines.

Although we were able to determine if written information on STIs was in line with BASHH PILs, there was variability in the ease of finding information between websites. Some pharmacies ensured that key messages were mandatory fields within the self-assessment questionnaire. Others buried key points within lots of text, which had the potential to be missed by patients.

One could argue that it would have been beneficial to have investigated STIs such as genital warts, scabies or pubic lice. However, we felt that current treatments available for these conditions were less likely to have an impact on antimicrobial resistance (AMR) which was the primary concern of this study. This is an area that we will look to explore in future work.

There were many positive exchanges with providers as a result of sharing our findings. This included meeting a medical advisor to an online pharmacy, an invitation to teach the headquarters of a market leading pharmacy and assurances that external agencies were supporting with written information.

Although our study is the first to investigate STI prescribing by online pharmacies, other studies have raised similar concerns about the ease of access to antibiotics online.[12]

## Conclusion

Patients want more autonomy over how they access Sexual Health with online assessment and postal treatment being acceptable to some patients. There is a paucity of data to explain why patients choose to engage with online pharmacies rather than Sexual Health services. One could surmise that online pharmacies offer flexibility compared to traditional “walk-in” Sexual Health clinics as patients can access care at a time and place that is convenient to them.

Prescribers on these platforms often have minimal expertise specific to Sexual Health. Our study has demonstrated that this leads to dubious advice regarding treatment, partner notification and STI testing. Harm reduction messaging e.g. chemsex support and HIV prevention was often missing.

Online pharmacies are not required to submit data to the UK Health Security Agency (UKHSA). This means STI surveillance is likely to be incorrect leading to delays in identifying outbreaks of infection and AMR. Although our results demonstrate we were able to improve standards, tougher regulation is needed to ensure that patients who choose to access online healthcare do not suffer as a consequence.

## Data Availability

All data including in this study was compiled into Excel spreadsheet format and is contained within the manuscript. Access to this data is available by contacting the lead author of this study.

